# A Network Analysis of Molecular Interactions to Study the Development of New-onset Diabetes and Hypertension after COVID-19 Infection Using Bioinformatics Tools

**DOI:** 10.1101/2023.09.10.23295323

**Authors:** Luis Jesuino de Oliveira Andrade, Luisa Correia Matos de Oliveira, Gabriela Correia Matos de Oliveira, Luís Matos de Oliveira

## Abstract

**Introduction:** The association between COVID-19 infection and the development of new-onset diabetes and hypertension is an emerging area of research. However, a comprehensive understanding of the underlying molecular mechanisms is still lacking. Network analysis using bioinformatics tools can provide valuable insights into the complex molecular interactions involved in these conditions after COVID-19 infection.

**Objective:** This study aims to use bioinformatics tools to analyze the network of molecular interactions related to new-onset diabetes and hypertension following COVID-19 infection.

**Methods:** Data from publicly available databases were utilized, including gene expression profiles and protein-protein interaction information. Differential expression analysis was performed to identify genes that were differentially expressed in individuals with new-onset diabetes and hypertension after COVID-19 infection compared to healthy controls. A protein interaction network was constructed using bioinformatics tools to explore the functional relationships among the identified differentially expressed genes.

**Results:** The network analysis revealed several key proteins and pathways related to the pathogenesis of new-onset diabetes and hypertension after COVID-19 infection. Notably, proteins involved in insulin signaling, glucose metabolism, inflammation, and blood pressure regulation were found to be prominently associated. The signaling pathway and the renin-angiotensin system were identified as key pathways in this context.

**Conclusion:** This study provides insights by showing a network-based perspective on the molecular interactions involved in the development of new-onset diabetes and hypertension after COVID-19 infection.

## INTRODUCTION

Diabetes and hypertension are two health conditions that continue to present significant challenges to public health worldwide.^1,2^ These diseases involve interactions of pathways making their understanding and management quite intricate.^3^

The COVID-19 pandemic caused by the severe acute respiratory syndrome coronavirus 2 has led to a global health crisis with wide-ranging implications. As research progresses, it is becoming evident that COVID-19 infection can have long-term impacts on various organ systems, including the development of new-onset diabetes and hypertension.^4^ The association between COVID-19 infection and the onset of these metabolic disorders has sparked significant interest among researchers worldwide, as understanding the underlying molecular mechanisms is crucial for effective management and prevention strategies.^5^

Emerging evidence suggests that COVID-19 infection can lead to dysregulation of glucose metabolism and blood pressure control, resulting in the manifestation of new-onset diabetes and hypertension in susceptible individuals.^4^ However, the precise molecular interactions responsible for this phenomenon remain poorly understood. In recent years, network analysis using bioinformatics tools has emerged as a powerful approach to unravel complex molecular interactions and pathways involved in various diseases.^6^

Several studies have already highlighted the potential role of specific pathways and molecules in the pathogenesis of new-onset diabetes and hypertension after COVID-19 infection. For instance, insulin signaling abnormalities and dysregulation of glucose metabolism has been implicated in COVID-19-associated diabetes development^4^. Moreover, inflammation, a hallmark feature of COVID-19 infection, has shown to play a crucial role in the pathogenesis of both hypertension and diabetes.^7^ Additionally, the renin-angiotensin system, which plays a crucial role in blood pressure regulation, has also been implicated in COVID-19-associated hypertension.^8^

The application of network analysis in the study of molecular interactions related to diabetes and hypertension offers numerous advantages. It allows researchers to identify key molecules and pathways involved in disease progression, providing potential biomarkers and therapeutic targets.^9^ Furthermore, network analysis can reveal hidden relationships between seemingly unrelated genes or proteins, shedding light on novel mechanisms underlying disease pathogenesis.^10^ By integrating information from different sources and incorporating various data mining algorithms, network analysis facilitates the discovery of molecular signatures associated with specific disease phenotypes, improving the accuracy of diagnosis and prognosis.

The network analysis using bioinformatics tools has revolutionized our ability to evaluate the intricate molecular interactions underlying diabetes and hypertension, and moving forward, continued advancements in network analysis methodologies and the incorporation of multi-omics datasets will be a great promise for improving the management and prevention of diabetes and hypertension. Thus, overall objective of this study is to evaluate the molecular mechanisms responsible for new-onset diabetes and hypertension following COVID-19 infection, using network analysis and bioinformatics tools.

## MATERIALS AND METHODS

### 1. Data Sources

Relevant datasets were collected from NCBI databases, including gene expression profiles and protein-protein interaction information. These datasets consisted of samples from individuals with new-onset diabetes and hypertension after COVID-19 infection.

### 2. Data Preprocessing

The collected datasets went through pre-processing to ensure data quality and consistency. This step included normalization of gene expression data, removal of any outliers or missing values, and standardization of data formats across different databases. Gene expression data were normalized using methods such as Reads Per Kilobase per Million mapped reads.

### 3. Integration of Datasets

The gene expression profiles and protein-protein interaction information were integrated to create a comprehensive dataset that captures the molecular interactions related to new-onset diabetes and hypertension following COVID-19 infection.

### 4. Network Construction

Bioinformatics tool such as STRING was used to construct a network model represented the molecular interactions identified in the integrated dataset. This network was providing a visual representation of the complex relationships among genes and proteins involved in these conditions.

### 5. Network Analysis

Various network analysis techniques, including network topology analysis and module identification, were applied to the constructed network. This analysis was helping identify key genes, proteins, and pathways that play a significant role in the development of new-onset diabetes and hypertension after COVID-19 infection.

### 6. Functional Annotation and Pathway Enrichment Analysis

The genes and proteins within the network were functionally annotated using databases and tools, such the Kyoto Encyclopedia of Genes and Genomes (KEGG).^11^ Pathway enrichment analysis was performed to determine the biological pathways that are significantly enriched in the molecular interactions associated with these conditions.

According to National Research Ethics Committee, the present study did not require submission to the Ethics Committee for evaluation because, as stated by the resolution, “the exemption from submission to the CEP/Conep System is limited to cases where the data is already provided in an aggregated form.”

## RESULTS

In total, 2,399 genes related to diabetes and 1,283 genes related to hypertension were found to construct the protein-protein interaction network of diabetes and hypertension with COVID-19. The data collected was pre-processed and a protein-protein interaction was established between 304 genes related to diabetes and COVID-19, and 211 genes related to hypertension and COVID-19.

The gene expression profiles and protein-protein interaction information are shown in intersection targets (Figure 1).

**Figure 1.**
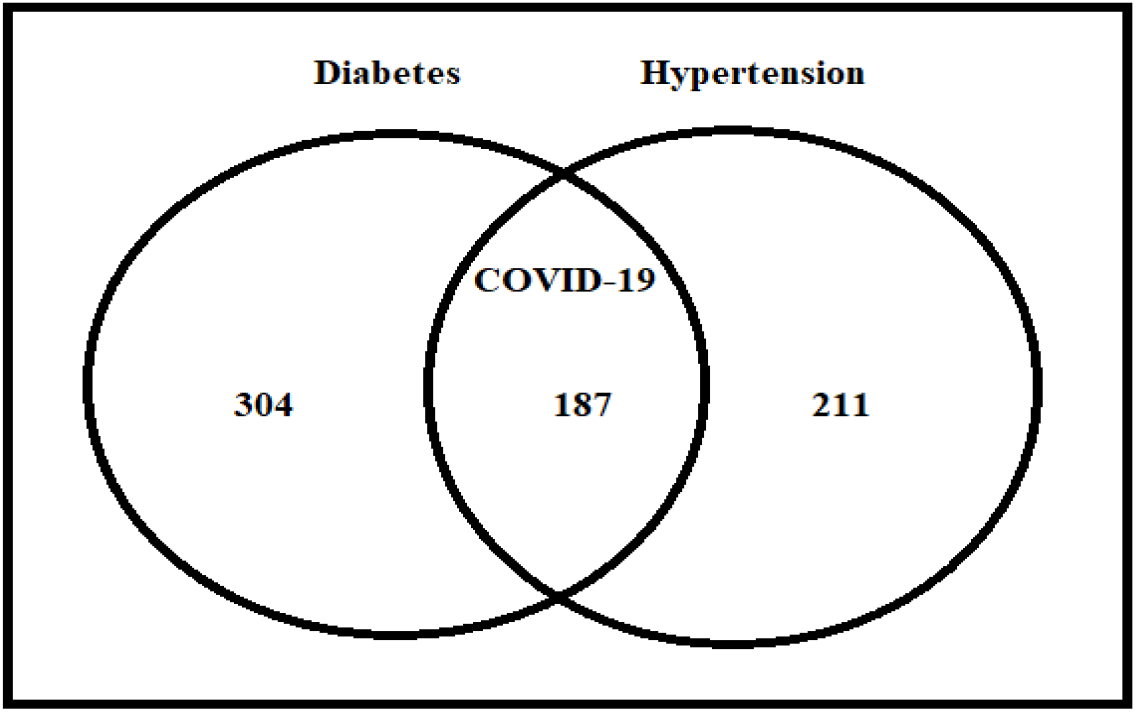
Interintersection of diabetes and hypertension overlap with COVID-19 **Source**: Survey results

The visual representation of the complex relationships between genes and proteins involved in these conditions is presented in **Figure 2**.

**Figure 2.**
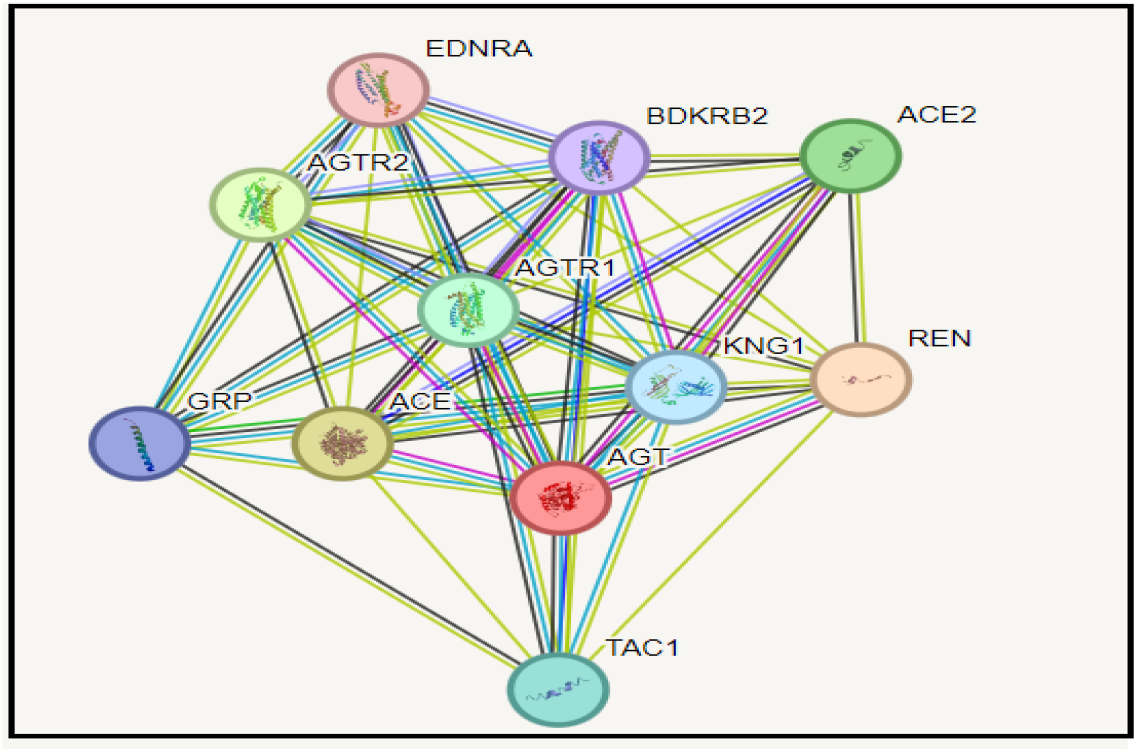
Relationships among diabetes, hypertension and COVID-19 genes **Source**:https://string-db.org/cgi/network?taskId=bO0JUIY9E5CJ&sessionId=biwngRTXo6TG

The main genes related to diabetes, hypertension, and COVID-19 that were found are as follows: the ACE2 (Angiotensin-converting enzyme 2) gene. The expression of ACE2 is upregulated in response to SARS-CoV-2 infection. Variations in this gene may influence susceptibility to COVID-19 and subsequent comorbidities. The SLC30A8 (Zinc transporter 8) gene, which has been associated with an increased risk of type 1 diabetes, and its expression may be influenced by SARS-CoV-2 infection. Additionally, the TCF7L2 (Transcription Factor 7-Like 2) gene has consistently been associated with an increased risk of type 2 diabetes.

The protein-protein interactions that are associated with the onset of new-onset diabetes and hypertension post-COVID-19 are as follows: cytokines, such as interleukin-6, and tumor necrosis factor-alpha, are released during COVID-19 infection and can lead to insulin resistance and hypertension. Additionally, dysregulation in the Renin-Angiotensin System pathway, including renin, ACE, and angiotensin II receptor type 1, can contribute to hypertension and diabetic complications.

The signaling pathways related to the onset of recent diabetes and hypertension post-COVID-19 is the following: the Insulin Signaling Pathway (Figure 3), where the disruption of insulin signaling due to inflammation and cytokine release leads to insulin resistance, hyperglycemia, and subsequent diabetes. Additionally, the Oxidative Stress Pathway (Figure 4) is relevant as COVID-19 infection induces oxidative stress, which contributes to the development of insulin resistance, vascular dysfunction, and endothelial damage leading to hypertension.

**Figure 3.**
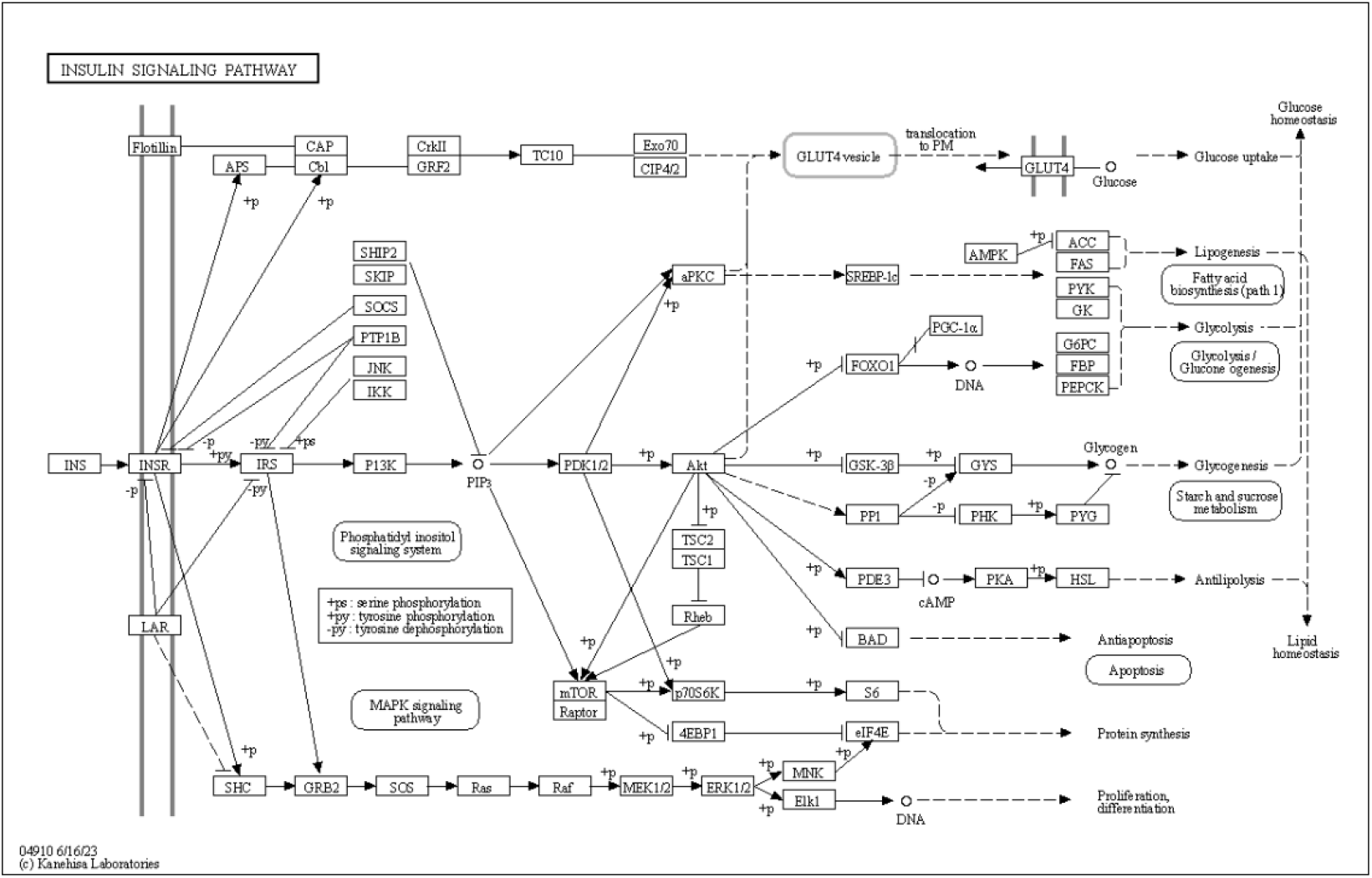
Insulin Signaling Pathway **Source**:https://www.kegg.jp/pathway/map=map04910&keyword=Insulin%20Signaling%20Pathway

**Figure 4.**
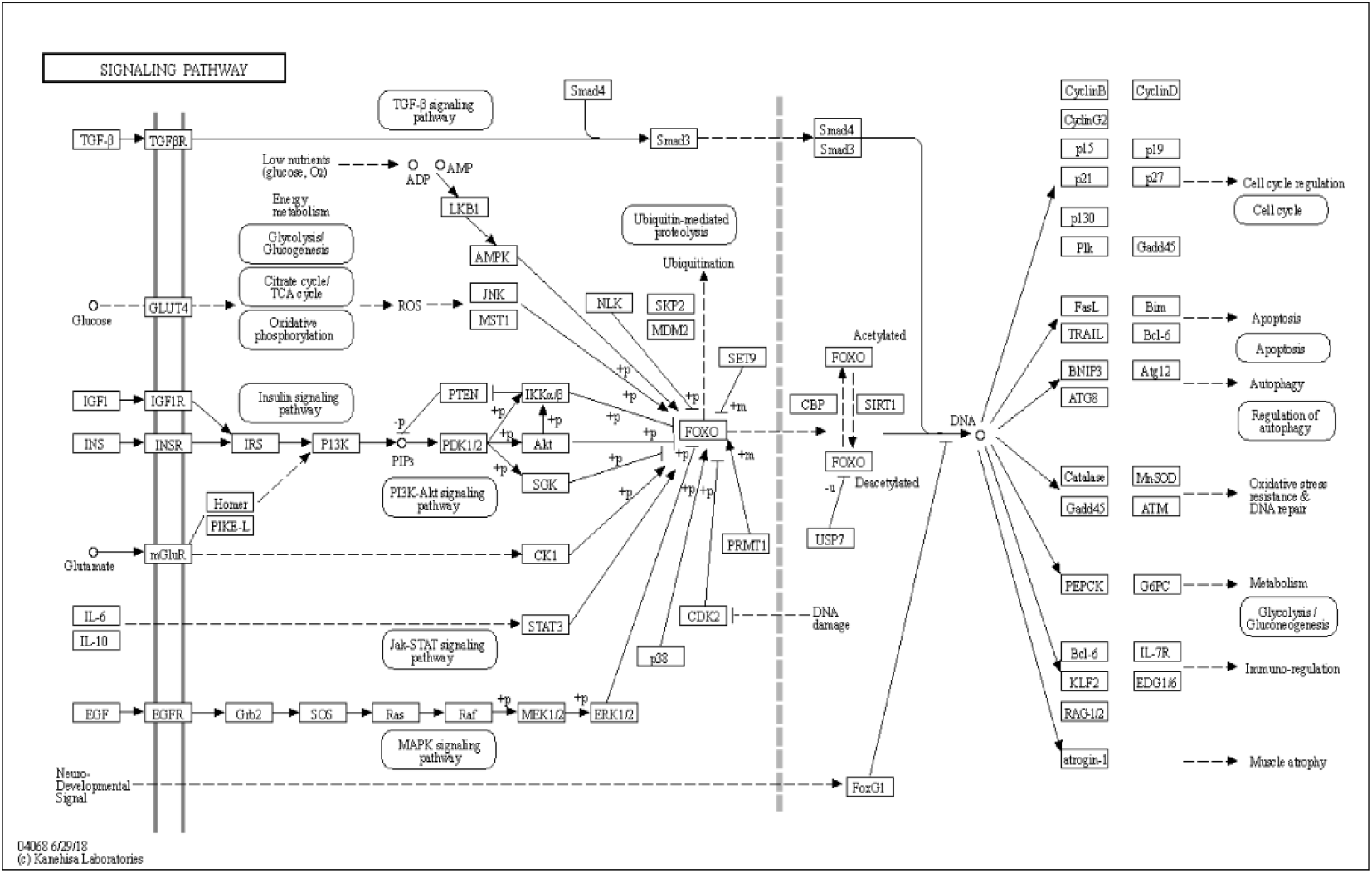
Oxidative Stress Pathway **Source**:https://www.kegg.jp/pathway/map=map04068&keyword=Oxidative%20Stress%20Pathway

The findings from the network analysis and pathway enrichment analysis were further validated and integrated with existing literature and experimental data. This step was enhancing the confidence in the identified molecular interactions and pathways related to new-onset diabetes and hypertension following COVID-19 infection.

## DISCUSSION

Our study demonstrated that the network analysis of molecular interactions can be used to study the development of new-onset diabetes and hypertension after a COVID-19 infection using bioinformatics tools.

Research has indicated that individuals who have recovered from COVID-19 face an increased likelihood of developing diabetes and hypertension. This heightened risk can be attributed to the physiological changes that occur during infection with the SARS-CoV-2 virus.^12,13^ Specifically, it is believed that pancreatic injury caused by the virus’s interaction with ACE2, a protein involved in regulating insulin secretion and glucose metabolism, plays a significant role in these metabolic alterations.^14,15^ Furthermore, studies have detected the presence of SARS-CoV-2 in pancreatic cells, suggesting that the virus may reprogram beta cells to produce glucagon, thereby triggering diabetes mellitus.^16^ In our research, we observed a strong association between the ACE2 gene and the SLC30A8 gene in relation to the response to SARS-CoV-2 infection. The ACE2 gene was found to have a positive regulatory effect on this response, while the SLC30A8 gene was found to be linked to an elevated risk of developing type 1 diabetes. Additionally, we identified the TCF7L2 gene, which has been previously associated with type 2 diabetes. These findings further contribute to our understanding of the genetic factors involved in diabetes susceptibility and the immune response to viral infections.

The Angiotensin system is implicated in the pathogenesis of COVID-19. Primarily, ACE2 functions as the cellular receptor for SARS-CoV-2, and the expression of the ACE2 gene could potentially regulate an individual’s susceptibility to infection.^17^ Additionally, ACE2 is considered a counterbalancing element in determining the risk of developing hypertension.^18^ Functional variations in the ACE2 genes have been associated with the risk of cardiovascular and pulmonary diseases, which could also potentially contribute to the outcome of COVID-19.^19^ Our study identified 211 genes that are associated with the connection between arterial hypertension and COVID-19, with the ACE2 gene being the primary gene involved in triggering post-COVID-19 hypertension.

It has been demonstrated the significance of understanding the protein-protein interactions involved diabetes and COVID-19.^20^ One such interaction of interest is the binding of the spike protein of SARS-CoV-2 to the ACE2 receptor, which is known to be regulated by the SLC30A8 gene. This interaction not only plays a crucial role in viral entry into cells but also raises concerns about the impact on glucose metabolism.^21^ Protein-protein interactions associated with the onset of diabetes and post-COVID-19 hypertension are observed, with cytokines such as interleukin-6 (IL-6) and tumor necrosis factor alpha (TNF-alpha) being released during COVID-19 infection.^22,23^ In our study, we identified protein-protein interactions associated with the onset of diabetes and post-COVID-19 hypertension involving IL-6, TNF-alpha, as well as renin, ACE, and angiotensin II.

Another significant factor in the initiation of diabetes and post-COVID-19 hypertension is the activation of signaling pathways, such as the Insulin Signaling Pathway and the Oxidative Stress Pathway,^24,25^ which play a crucial role in the development of insulin resistance, vascular dysfunction, and endothelial damage ultimately leading to hypertension.^26^ In our study, we substantiated these pathways by referencing KEGG^10^ as a point of reference.

Thus, in the present study we evaluated the molecular interactions involved in the development of recently onset diabetes and hypertension after COVID-19 infection through a comprehensive network analysis using bioinformatics tools. By analyzing the intricate connections between various molecules, such as proteins, genes, and metabolites, we assessed the underlying mechanisms responsible for the occurrence of these post-COVID-19 complications. By performing the network analysis, which involved the integration of multiomic data, we obtained an understanding of the molecular alterations occurring in individuals with recently onset diabetes and hypertension post-COVID-19.

## Data Availability

All data produced in the present work are contained in the manuscript

